# Procalcitonin to Predict Severity of Acute Cholangitis and Need for Urgent Biliary Decompression: Systematic Scoping Review

**DOI:** 10.1101/2021.08.24.21262522

**Authors:** Krixie Silangcruz, Yoshito Nishimura, Torrey Czech, Nobuhiko Kimura, James Yess

## Abstract

**Background:** Serum Procalcitonin (PCT) has been reported as a potential biomarker to predict the severity of acute cholangitis (AC) or the need for urgent biliary decompression. This study aimed to identify and summarize the existing research about the serum PCT and the severity of AC, and to find gaps where future studies can be targeted towards.

**Methods:** Following the PRISMA Extension for Scoping Reviews, MEDLINE, EMBASE, and Google Scholar were searched for all peer-reviewed articles with relevant keywords including “cholangitis” and “procalcitonin” from their inception to July 13, 2021.

**Results:** We identified six studies. All the studies employed case-control design and aimed to evaluate the usefulness of serum PCT to predict the severity of AC with key identified outcomes. While potential cut-off values of serum PCT for severe AC ranged from 1.8–3.1 ng/mL, studies used different severity criteria and the definition of urgent biliary decompression. No studies proposed cut-off PCT values for the need for urgent biliary decompression.

**Conclusion:** This scoping review identified that the current level of evidence regarding the usefulness of serum PCT in assessing the severity of AC. Further clinical research is warranted with a focus on standardized outcome measures employing prospective or experimental designs.

## Introduction

Acute cholangitis (AC) is a medical emergency and systemic condition due to biliary infection and obstruction with an associated high mortality rate^1, 2^. Before advancements in critical care and decompression of the biliary duct system, the mortality of AC was reported to be over 50%^2^. Subsequent to 1980, the mortality rates of AC ranged from 10-30%, with multiorgan failure noted to be the cause of death^3^. The majority of cases of AC are due to biliary duct stones. The variety of additional etiologies underscores a wide variety of risk factors that influence mortality^3, 4^.

Procalcitonin (PCT), a 116 amino acid peptide precursor of calcitonin, is initially thought to help identify sepsis patients and was later associated with bacterial infection^5^. However, its role in disease severity prediction remains unclear^6^, with a reported sensitivity and specificity of PCT to predict septic shock of any causes 63% and 65%, respectively^7^. Thus, the use of PCT in the diagnosis of sepsis remains a topic of contentious debate^8^.

Various studies have found PCT to be correlated with the disease severity of AC. The Tokyo Guidelines 2018 (TG18) for AC discussed the utility of PCT as a parameter for the severity assessment, stating that there was level D (very low-quality) evidence, and it might be an area of future research^9^. So far, small-scale studies have been conducted to examine the usefulness of serum PCT values to predict the severity of AC or the need for urgent biliary decompression, using either Tokyo Guidelines 2007 (TG07) and Tokyo Guidelines 2013 (TG13)^10-15^. However, to date, no systematic reviews are available to analyze the evidence from studies across different settings and clinical outcomes to identify the trends of PCT in AC or limitations with current literature to generate recommendations in this area for future research.

The objective of this study is to scope the states of research to determine the relationship between serum PCT levels and AC disease severity, trends correlating PCT levels to different hierarchical levels of intervention as well as the need for urgent biliary decompression to identify directions for future research in this area.

## Methods

### Study Design

This is a systematic scoping review conducted in accordance with the Preferred Reporting Items for Systematic Reviews and Meta-Analyses (PRISMA) extension for scoping reviews (PRISMA-ScR)^16, 17^. See **Appendix 1** for PRISMA-ScR Checklist of the present study.

### Search Strategy

We searched MEDLINE, EMBASE, and Google Scholar for all peer-reviewed articles and conference abstracts from inception to July 13^th^, 2021. No filters for study design and language were used. A manual screening for additional pertinent articles was done using the reference lists of all articles that met the eligibility criteria. The search strategy involved relevant keywords, including “cholangitis” and “procalcitonin.” The search was conducted by two authors (YN and NK) independently. See **Appendix 2** for detailed search terms.

### Eligibility Criteria

The criteria for the inclusion of articles are the following:

1. Peer-reviewed articles evaluating the relationship between serum procalcitonin levels and severity of acute cholangitis or need for biliary decompression.
2. Randomized controlled trials (RCTs), case-control studies, cohort studies (prospective or retrospective), cross-sectional studies, and case series in adult patients

The exclusion criteria included the following:

1. Qualitative studies, review articles, case reports, and commentaries.
2. Conference abstracts
3. Studies involving pediatric or obstetric patients.

### Study Selection

Articles selected for full-text assessment were assessed independently by YN and NK using EndNote 20 reference management software. Articles considered eligible were then evaluated in full length with the inclusion and exclusion criteria.

### Data Extraction

A standardized data collection form that followed the PRISMA and Cochrane Collaboration guidelines for systematic reviews was used to obtain the following information from each study: title, name of authors, year of publication, country of origin, study characteristics, target outcome, aims, study and comparative groups, key findings, and limitations.

## Results

### Search Results and Study Selection

**Figure 1.** shows a PRISMA flow diagram that depicts the process of identification, screening, eligibility, and inclusion or exclusion of the studies. The initial search of MEDLINE, EMBASE and Google Scholar yielded 1987 articles. 62 duplicate studies were removed, followed by the elimination of 1900 articles that were either irrelevant to our study, review articles, editorial articles, or conference abstracts with title and abstract screening. Subsequently, 25 articles underwent a full-text review. Of these articles, 19 articles were because they were review articles, did not investigate the need for biliary drainage or cholangitis severity as an outcome, did not measure procalcitonin in the participants, or were not studies about procalcitonin and acute cholangitis. One study was also excluded because of the discrepancy between the data presented in the main text and the figures.

**Figure 1.**
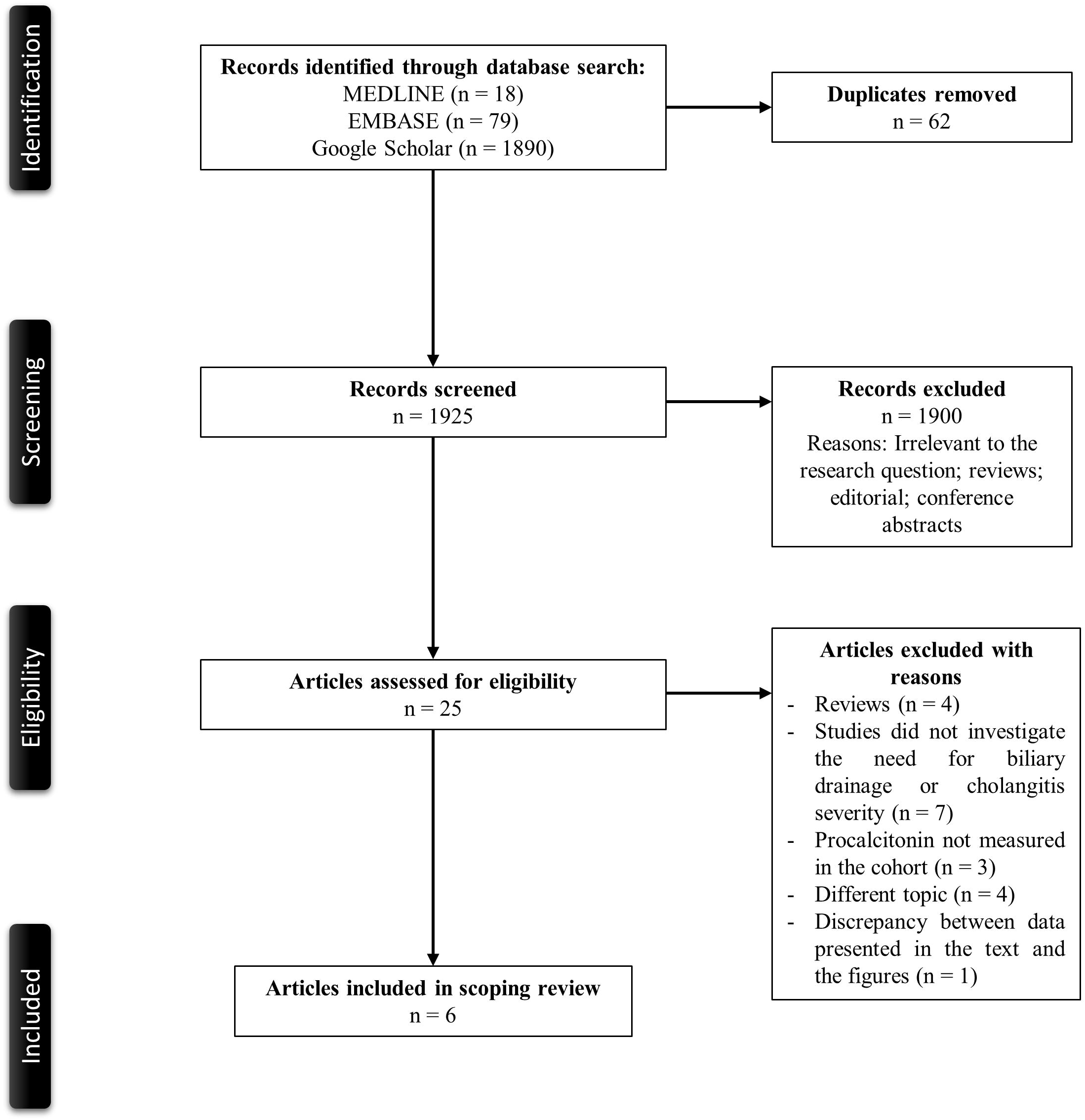
PRISMA flowchart of the search strategy.

### Description of Included Studies

A total of six studies met our eligibility criteria for the scoping review^10-12, 14, 18, 19^. The main characteristics of the included studies are described in **Table 1**. Five were retrospective case-control studies, while one study^12^ used a prospective case-control design. All the studies were from Asia (Japan, n = 4; South Korea, n =1; China. n = 1). Sample sizes varied from 58 to 213 participants. All studies aimed to evaluate the usefulness of PCT to predict the severity of AC and were based on data from a single center.

**Table 1.**
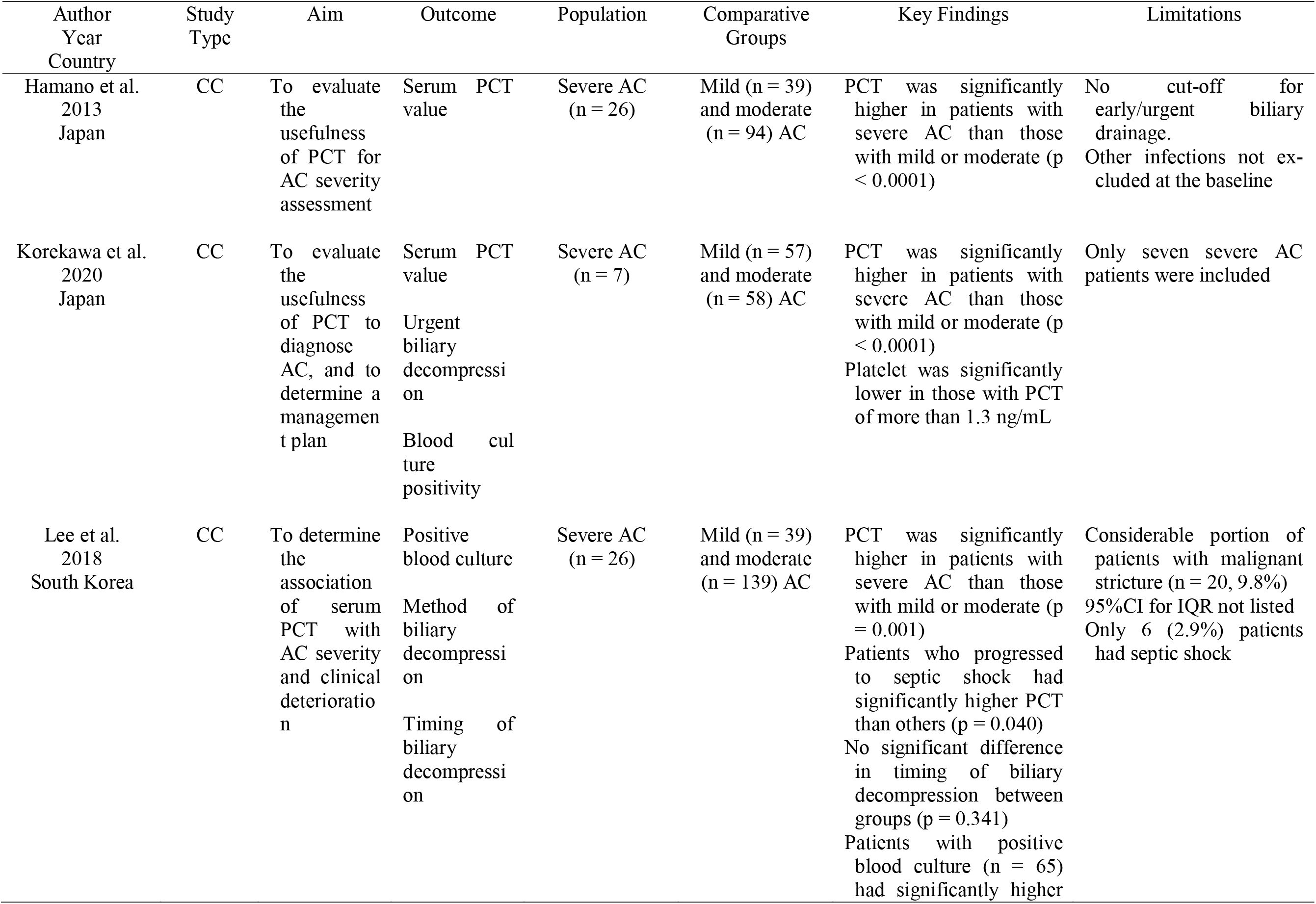
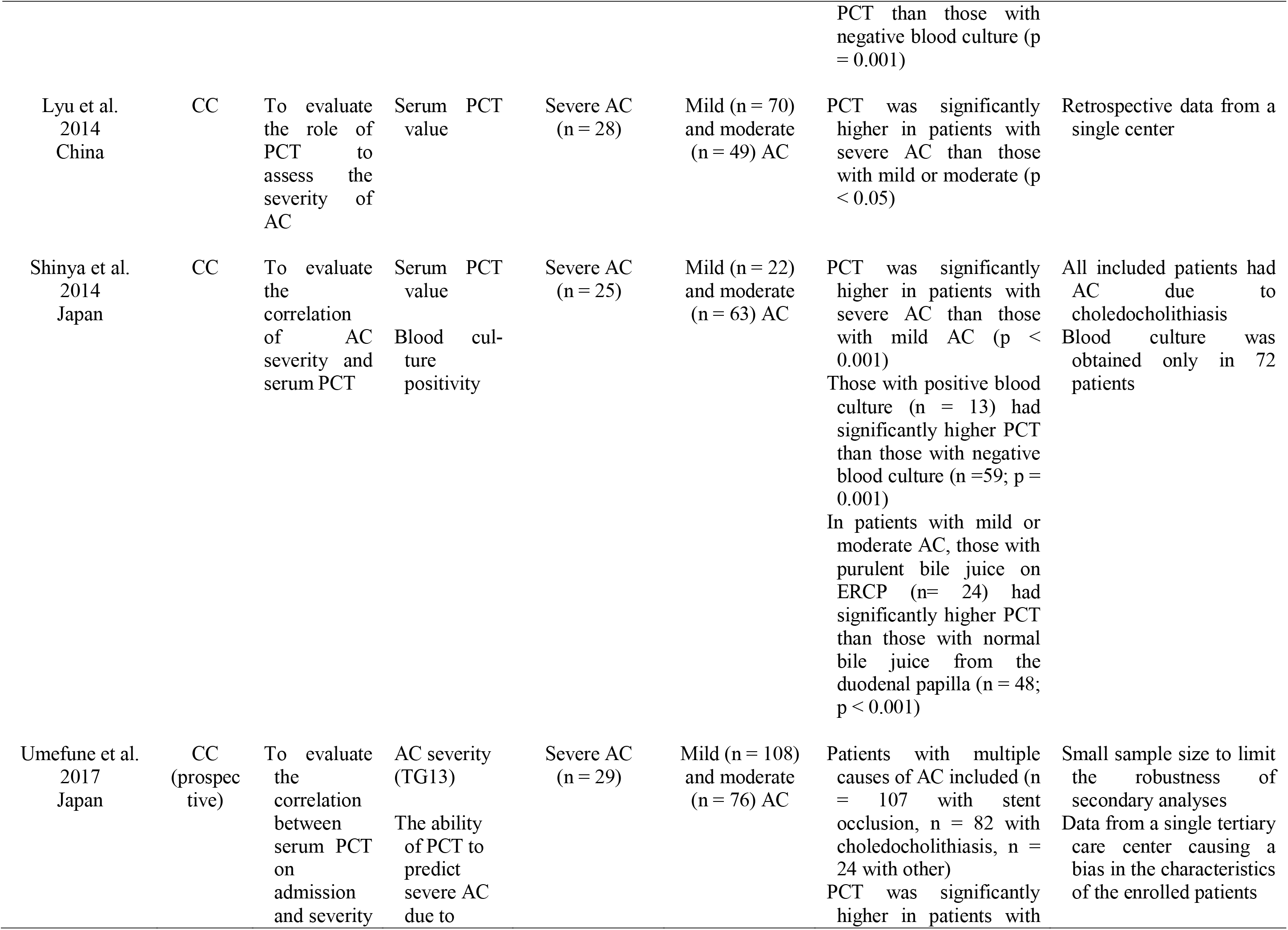
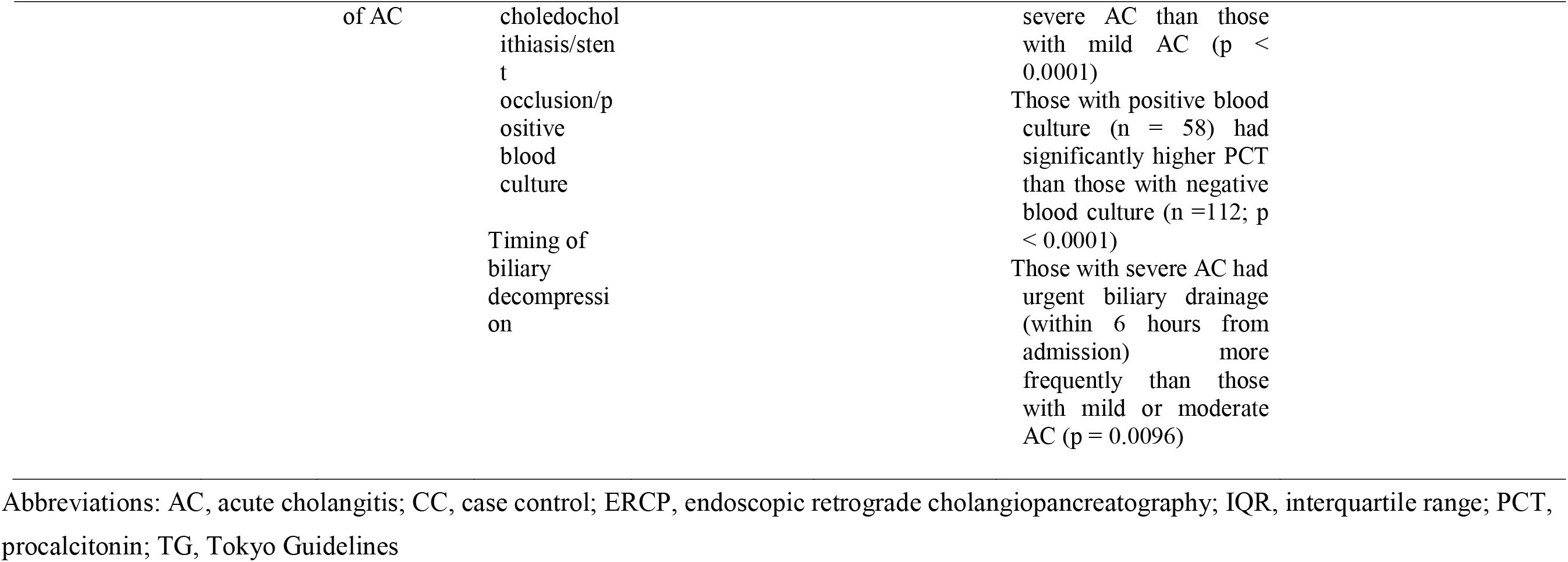
Main characteristics of the included studies in the scoping reviews.

### Outcomes

Four studies also assessed blood culture positivity among participants^11, 12, 14, 19^. The number of patients who went through urgent biliary decompression was reported in three studies^12, 14, 19^. Of the three studies, a study by Lee et al. and Umefune et al. reported timing of biliary decompression^12, 14^. 4 studies assessed positivity of blood culture in patients with AC^11, 12, 14, 19^.

### Severity of Acute Cholangitis

Findings related to serum PCT levels and the severity of AC or other pertinent outcomes are summarized in **Table 2**. Except for one study^10^, TG13 Guidelines were used as the Severity Criteria of AC. There was a significant variation in the proportion of patients with severe AC from 7/122 (5.7%)^19^ to 25/110 (22.7%)^11^. All studies reported that serum PCT levels were significantly higher in those with severe AC than those with mild or moderate AC, although the median serum PCT level differed considerably among the included articles. Area under the receiver operator characteristic (AU-ROC) of serum PCT for severe AC varied from 0.75 (95% confidence interval [CI] 0.63–0.87)^11^ from 0.90 (95%CI 0.85–0.96)^12^. Except for one study^19^, potential cut-off values of serum PCT for severe AC were proposed, which ranged from 1.76 ng/mL (sensitivity 84.6%, specificity 62.4%) from 3.1 ng/mL (sensitivity 80.8%, specificity 84.6%)^10^ using the Youden Index method, with three studies that reported similar cut-off values (Umefune et al. 2.2 ng/mL, Shinya et al. 2.33 ng/mL, Lyu et al. 2.38 ng/mL).

**Table 2.** Key findings related to serum procalcitonin levels in the included studies.

| Author<br>Year<br>Country | Severity<br>Criteria<br>Used | PCT – median (ng/mL) |  |  | Cut-off PCT for urgent<br>biliary decompression<br>(ng/mL) | Key Findings |
| --- | --- | --- | --- | --- | --- | --- |
|  |  | Mild AC | Moderate AC | Severe AC |  |  |
| Hamano et al.<br>2013<br>Japan | TG07 | 0.08<br>(IQR;<br>0.04–0.18) | 0.37<br>(IQR;<br>0.15–1.85) | 5.56<br>(IQR;<br>3.59–25.89) | N/A | Cut-off PCT for severe AC: 3.1 ng/mL<br>(sensitivity 80.8%, specificity 84.6%, OR 23.1<br>[95% CI 8.0–70.2])<br>AU-ROC of PCT for severe vs. mild to<br>moderate AC = 0.86 (95% CI 0.78–0.92) |
| Korekawa et al.<br>2020<br>Japan | TG13 | 0.9<br>(IQR;<br>0.1–1.1) | 9.9<br>(IQR;<br>1.2–15.9) | 37.8<br>(IQR;<br>20.3–54.9) | N/A<br>(cut-off PCT to predict<br>those with lower<br>platelet, who might be<br>complicated with DIC<br>and potential<br>candidates for urgent<br>biliary decompression) | AU-ROC of PCT for severe and moderate vs.<br>mild AC = 0.89 (95% CI 0.84–0.95); no cut-off<br>value provided<br>Median PCT of those with positive blood<br>culture: 12.7 ng/mL (IQR, 0.45–19.8) vs. 4.6<br>ng/mL (IQR, 0.2–4.9) in those with negative<br>blood culture<br>Cut-off PCT for positive blood culture: 1.3<br>ng/mL (sensitivity 67.0%, specificity 55.0%)<br>Those with PCT > 1.3 ng/mL had significantly<br>lower platelet than others (p < 0.0001) |
| Lee et al.<br>2018<br>South Korea | TG13 | 0.22<br>(IQR; 0.52) | 1.35<br>(IQR; 4.67) | 9.41<br>(IQR, 53.69) | N/A | Median PCT of those with positive blood<br>culture: 3.25 ng/mL (IQR, 8.86) vs. 0.62<br>ng/mL (IQR, 3.78) in those with negative<br>blood culture<br>Median PCT of those progressed to septic<br>shock: 9.11 ng/mL (IQR, 18.52) vs. 0.89<br>ng/mL (IQR, 4.34) in others<br>Cut-off PCT for severe AC / septic shock: 1.76<br>ng/mL (sensitivity 84.6%, specificity 62.4%) /<br>3.77 ng/mL (sensitivity 80.0%, specificity<br>74.0%) |
|  |  |  |  |  |  | AU-ROC of PCT for severe vs. mild to moderate AC = 0.778 (95%CI 0.680–0.876) |
| Lyu et al.<br>2014<br>China | TG13 | 0.166 (mean;<br>SD ± 0.033) | 0.349 (mean;<br>SD ± 0.046) | 0.759 (mean;<br>SD ± 0.029) | N/A | Cut-off PCT for severe AC / moderate AC: 2.38 ng/mL (sensitivity 78.9%, specificity 73.7%) / 0.162 ng/mL (sensitivity 75.0%, specificity 61.1%) |
| Shinya et al.<br>2014<br>Japan | TG13 | 0.2 (IQR;<br>0.1–0.7) | 0.7 (IQR;<br>0.2–2.7) | 6.8 (IQR;<br>0.5–48.3) | N/A<br>(cut-off PCT to predict those with purulent bile juice from the duodenal papilla was noted) | Cut-off PCT for severe AC: 2.33 ng/mL (sensitivity 64.0%, specificity 78.0%)<br>AU-ROC of PCT for severe AC = 0.75 (95%CI 0.63–0.87)<br>Median PCT of those with purulent bile juice: 7.9 ng/mL (IQR; 1.2–37.7) vs. 0.6 (IQR; 0.2–2.9) in those with normal bile juice<br>Cut-off PCT for purulent bile juice: 3.2 ng/mL (sensitivity 67.0%, specificity 79.0%)<br>AU-ROC of PCT for purulent bile juice = 0.77 (95%CI 0.64–0.89) |
| Umefune et al.<br>2017<br>Japan | TG13 | 0.45<br>(IQR;<br>0.22–1.69) | 1.25<br>(IQR;<br>0.41–4.18) | 19.51<br>(IQR;<br>4.41–53.36) | N/A | Cut-off PCT for severe AC: 2.2 ng/mL (sensitivity 97.0%, specificity 73.0%)<br>AU-ROC of PCT for severe AC = 0.90 (95%CI 0.85–0.96); better than WBC or CRP<br>Median PCT of those with positive blood culture: 4.71 ng/mL (IQR, 0.87–16.96) vs. 0.65 ng/mL (IQR, 0.25–2.26) in those with negative blood culture |

Abbreviations: AC, acute cholangitis; AU-ROC, area under the receiver operator characteristic; CI, confidence interval; CRP, C-reactive protein; DIC, disseminated intravascular coagulopathy; IQR, interquartile range; OR, odds ratio; PCT, procalcitonin; SD, standard deviation; TG, Tokyo Guidelines; WBC, white blood cell

### Procalcitonin and Urgent Biliary Decompression

Although some studies included the term “biliary drainage” or “biliary decompression”^11, 14^, no studies proposed potential cut-off values for the need for urgent or emergent biliary decompression. The surrogate of the need for urgent or emergent biliary decompression varied considerably among the studies. Korekawa et al.^11^ used thrombocytopenia as the surrogate and suggested that the cut-off serum PCT to predict lower platelet counts (1.3 ng/mL) might also be helpful to determine those who may need urgent biliary decompression. Shinya et al.^11^ proposed a cut-off PCT for the presence of purulent bile juice on biliary decompression (3.2ng/mL) as the surrogate. Other studies used the severity of AC as the surrogate for the need for urgent biliary decompression. Of note, the definitions of urgent or emergent decompression were different among the articles. Korekawa et al. defined “emergent endoscopic retrograde cholangiopancreatography (ERCP)” as “the procedures done within 24 hours after admission”, while Umefune et al. defined urgent and early biliary drainage as “biliary drainage six hours and 12 hours after admission”, respectively. Other studies did not specify the term of early, urgent, or emergent biliary decompression.

## Discussion

In this scoping review, we identified six primary studies related to serum PCT levels and the severity of AC or other pertinent outcomes such as blood culture positivity, progression to septic shock, need, and timing of biliary decompression, or biliary fluid character on ERCP. All studies reported that serum PCT levels were significantly higher in those with severe AC than those with mild or moderate diseases, although the median serum PCT levels were considerably different among the included articles.

AC requires appropriate treatment in the early phase because severe AC may result in death if no early appropriate medical care is provided. Historically, TG13 proposed management bundles of AC, and TG18 re-defined the management bundles and mentioned the potential utility of serum PCT in predicting the severity of AC^15, 20^. These guidelines classified the severity of AC into three grades; mild (grade I), moderate (grade II), and severe (grade III)^21^. In particular, TG18 severity grading criteria for AC are essential for predicting prognosis and determining a treatment strategy by identifying patients requiring early biliary drainage, although its ability to identify those who need the procedure is limited^22^. As noted above, there was very low-quality evidence suggesting the utility of PCT as a parameter for the severity assessment of AC when TG18 was proposed. Despite a suggestion that PCT might be a better biomarker to predict the severity of AC than white blood cell or C-reactive protein^12^, unfortunately, however, there is still limited evidence regarding the point, although three years have passed since TG18.

The present study results showed that previous literature had a certain degree of consistency regarding a potential serum PCT cut-off value for severe AC (ranged from approximately 1.8 ng/mL to 3.1 ng/mL) with satisfactory AU-ROC. However, all the studies had different limitations, as described in **Table 1**. While the proposed serum PCT cut-off values for severe AC were similar, median PCT values of either moderate or severe AC differed considerably among the included studies. Of note, no studies proposed cut-off values to proceed with urgent biliary decompression, and the severity of AC was used as a surrogate of the need for the procedure. Of note, there were inconsistencies in the severity criteria used (Hamano et al. used TG07 and the others employed TG13) and the definition of either “urgent” or “emergent” decompression. For example, Umefune et al. defined urgent biliary drainage as “within 6 hours from admission”, while Lee et al. classified patients according to the timing of biliary drainage; within 24 hours, from 24 to 48 hours, or after 48 hours from a hospital visit. The differences in the guidelines used and the outcome measurements negatively affect the level of evidence.

Other limitations of the studies include that all of the currently available research were case-control studies (five retrospective and one prospective) with small sample sizes. Also, considerable heterogeneity of basic demographics among the studies needs to be noted. All the studies were conducted in Asian countries, including Japan, South Korea, and China. Further, the etiology of AC may be different; for instance, 9.8% of patients in the study by Lee et al. had malignant biliary strictures, while Shinya et al. included only patients with choledocholithiasis. All the factors limit the generalizability of the results. Future studies may need adequate power with a prospective design and standardized outcome measurements, using the same severity criteria and the definition of urgent biliary decompression. Non-randomized controlled trial comparing the outcome of those with or without urgent biliary decompression might be necessary to establish a cut-off serum PCT value to determine the need for the procedure.

Several limitations in this study should be noted. First, due to the urgent need for evidence on this topic and limited time, we did not contact authors to clarify the details of data described in literature. Second, we only included peer-reviewed articles in this scoping review. Thus, non-peer-reviewed articles or conference abstracts, which might be useful, were excluded from the study. To the best of our knowledge, however, there has been no scoping review to investigate the utility of serum PCT to predict the severity of AC or the need for biliary decompression.

In conclusion, this scoping review identified that the current body of evidence regarding the usefulness of serum PCT in assessing the severity of AC, and potential cut-off values for the need for urgent biliary decompression, remains scarce. In the future, further clinical research employing prospective or experimental designs is imperative to determine practical cut-off values for serum PCT for severe AC and the need for early biliary decompression.

## Data Availability

Data is available on request from the authors.

## Conflicts of Interest

The authors declare no conflicts of interest in association with the present study.

## Funding

None.

## Author contributions

KS conceived the study and drafted the manuscript. YN searched the literature, assessed the quality of the studies, drafted and revised the manuscript. TC performed data interpretation, drafted and revised the manuscript. NK searched the literature and revised the manuscript. JY revised the manuscript.

## Notes

### Competing Interest Statement

The authors have declared no competing interest.

