## Appendix 2 for "Procalcitonin to Predict Severity of Acute Cholangitis and Need for Urgent Biliary Decompression: Systematic Scoping Review"

Medline

(“cholangitis”[MeSH Terms] OR “Cholangitis”[All Fields]) AND (“procalcitonin”[MeSH Terms] OR “Procalcitonin”[All Fields])

EMBASE

(‘cholangitis’/exp OR cholangitis) AND (‘procalcitonin’/exp OR procalcitonin)

Google Scholar

Cholangitis AND Procalcitonin
