## Appendix 3 for "Procalcitonin to Predict Severity of Acute Cholangitis and Need for Urgent Biliary Decompression: Systematic Scoping Review"

**TG18/TG13 Severity Grading of Acute Cholangitis**

| **Grading** | | |
| --- | --- | --- |
| (A) Cardiovascular dysfunction  Hypotension requiring dopamine (≥5 μg/kg per min or any dose of norepinephrine) | Yes | No |
| (A) Neurological dysfunction: Disturbance of consciousness | Yes | No |
| (A) Respiratory dysfunction (PaO₂/FiO₂ ratio <300) | Yes | No |
| (A) Renal dysfunction: Oliguria or creatinine >2.0 mg/dL | Yes | No |
| (A) Hepatic dysfunction (INR >1.5) | Yes | No |
| (A) Hematological dysfunction (Platelet count <100,000/mm³) | Yes | No |
| (B) Abnormal WBC count (>12,000/mm³ or <4,000/mm³) | Yes | No |
| (B) High fever (≥39°C/102.2°F) | Yes | No |
| (B) Age ≥75 years | Yes | No |
| (B) Hyperbilirubinemia (Total bilirubin ≥5 mg/dL) | Yes | No |
| (B) Hypoalbuminemia (<0.7 x upper limit of normal) | Yes | No |
| **Grade I Mild acute cholangitis: Not meeting the criteria for “severe” or “moderate”**  Recommendation: antibiotics and general supportive care; consider biliary drainage if no response to initial treatment | | |
| **Grade II Moderate acute cholangitis: Any two of the “B” criteria**  Recommendation: antibiotics and general supportive care; early endoscopic or percutaneous transhepatic biliary drainage is indicated | | |
| **Grade III Severe acute cholangitis: At least one of the “A” criteria**  Recommendation: initial treatment with antibiotics, urgent biliary drainage, appropriate respiratory/circulatory management | | |
